# Differences in Genetic Correlations between Posttraumatic Stress Disorder and Alcohol Use Disorder-Related Phenotypes Compared to Alcohol Consumption-Related Phenotypes

**DOI:** 10.1101/2022.03.02.22271415

**Authors:** Kaitlin E. Bountress, Daniel Bustamante, Stacey Subbie-Saenz de Viteri, Chris Chatzinakos, Christina Sheerin, Roseann E. Peterson, Bradley T. Webb, Nikolaos P. Daskalakis, Howard Edenberg, The Psychiatric Genomics Consortium Posttraumatic Stress Disorder Working Group, Jackie Meyers, Ananda Amstadter

## Abstract

**Background:** Posttraumatic Stress Disorder (PTSD) tends to co-occur with greater alcohol consumption as well as alcohol use disorder (AUD). However, it is unknown whether the same etiologic factors that underlie PTSD-AUD comorbidity also contribute to PTSD-alcohol consumption.

**Methods:** We used summary statistics from large-scale genome-wide association studies (GWAS) of European-ancestry (EA) and African-ancestry (AA) participants to estimate genetic correlations between PTSD (both the diagnosis and re-experiencing symptoms) and a range of alcohol consumption-related and AUD-related phenotypes (e.g., drinks per week, max drinks, consumption, AUD).

**Results:** In EAs, there were positive genetic correlations between PTSD phenotypes and AUD-related phenotypes (i.e., Alcohol Use Disorders Identification Test (AUDIT) problem score, maximum alcohol intake, AUD, and alcohol dependence) (rGs: .132-.533, all FDR adjusted p<.05). However, the genetic correlations between PTSD phenotypes and alcohol consumption -related phenotypes (i.e., drinks per week, AUDIT consumption score, AUDIT total score, and a combination of consumption and problems) were negatively associated or non-significant (rGs: -.417- -.042, FDR adjusted p: <.05-NS). For AAs, the direction of correlations was sometimes consistent and sometimes inconsistent with that in EAs, and the ranges were larger (rGs for AUD--related: -.275 -.266, FDR adjusted p: NS, alcohol consumption-related: .145-.699, FDR adjusted p: NS).

**Conclusions:** These findings illustrate that the genetic associations between consumption and problem alcohol phenotypes and PTSD differ in both strength and direction. Thus, the genetic factors that may lead someone to develop PTSD and consume large quantities of alcohol are not the same as those that lead someone to develop PTSD-AUD.

## Introduction

Traumatic events are common, with 50-70% of individuals experiencing at least one trauma in their lifetimes (Benjet et al., 2016). Posttraumatic Stress Disorder (PTSD), the signature trauma-related disorder (Breslau, 2009), is associated with increased alcohol consumption (Vlahov et al., 2002) and alcohol use disorder (AUD) (Jakupcak et al., 2010). Twin studies (Heath, Jardine, & Martin, 1989; Kaprio et al., 1987; Knopik et al., 2004; Stein, Jang,Taylor, Vernon, & Livesley, 2002) and genome-wide association studies (GWAS) (Clarke, Adams, Davies, Howard, Hall, Padmanabhan, Murray, Smith, Campbell, Hayward, Porteous, Deary, & McIntosh, 2017; Stein et al., 2016) find that PTSD and these alcohol phenotypes are moderately heritable, with 36-60% of the variance explained by genetic effects. Additional work using twin studies has demonstrated 30% genetic overlap between PTSD and AUD (McLeod et al., 2001; Xian et al., 2000). In general, most of the comorbidity research has focused on PTSD-AUD, and has neglected the association between PTSD-alcohol consumption. As increased alcohol consumption is associated with AUD (Moos, Schutte, Brennan, & Moos, 2004; Sanchez-Roige et al., 2019), genetic research is needed to test whether the same genetic influences underlying PTSD-AUD are those underlying PTSD and alcohol consumption.

Large scale GWAS have identified significant hits for PTSD phenotypes (e.g., Nievergelt et al., 2019), problematic alcohol use (PAU) (Zhou et al., 2020), (e.g., alcohol dependence; Walters et al., 2018), and alcohol consumption (T. K. Clarke et al., 2017). Recent analyses have allowed for examination of genetic associations across alcohol consumption (Kranzler et al., 2019; Liu et al., 2019; Sanchez-Roige et al., 2019), PAU (Gelernter, Sun, Polimanti, Pietrzak, Levey, Lu, et al., 2019; Sanchez-Roige et al., 2019; Walters et al., 2018; Zhou et al., 2020), as well as PTSD and re-experiencing symptoms (Gelernter, Sun, Polimanti, Pietrzak, Levey, Bryois, et al., 2019; Nievergelt et al., 2019). Single nucleotide polymorphism (SNP) -based heritability of PTSD suggest modest to moderate heritability (∼15%), with these estimates larger in women than men (i.e., 36% versus 5%; Duncan et al., 2017). SNP-based heritability of problem alcohol phenotypes suggests modest heritability, ranging from 5.6%-9.4% for AD (h2=.090, s.e.=.019; Walters et al., 2018), AUD (Kranzler h2=.056, s.e.=.004; Zhou h2=.094, s.e.=.005; Kranzler et al., 2019; H. Zhou et al., 2020), and PAU (h2=.068, s.e.=.004; H. Zhou et al., 2020).

Few have investigated the genetic association between PTSD and AUD using genetic techniques such as linkage disequilibrium score regression (Bulik-Sullivan et al., 2015), but work by our group found a significant correlation between PTSD and alcohol dependence (AD; rG = .35, Sheerin et al., 2020) (rG = .28; Bountress et al., 2021) for those of European Ancestry. However, this effect was driven by women, for whom the genetic correlation was moderate and significant, but not for men (Sheerin et al., 2020). Genetic correlation analyses between PTSD and alcohol consumption were also conducted by our group, finding a non-significant association (rG = -.07; Bountress et al., 2021); another group found near zero genetic correlation between PTSD and the Alcohol Use Disorders Identification Test (AUDIT) consumption score subscale (AUDIT-C)(Mallard et al., 2021). Work by our group also found that beyond genetic correlations, using Mendelian Randomization, PTSD exerted a causal effect on AUD, but not alcohol consumption, but that neither alcohol phenotype exerted a causal influence on PTSD (Bountress et al., 2021). Additionally, genetic correlations between consumption and problems phenotypes vary. One group found correlations between alcohol consumption and AUD ranging from small to moderate (e.g., ∼rG=.2-.3; Sanchez-Roige et al., 2019). Another found large associations between AUDIT-C and AUDIT-P and AD (∼ rG=.70; Mallard et al., 2021) once the association between the frequency item and SES was taken into account. Together these findings suggest the genetic risk for consumption and problematic phenotypes are correlated but distinct.

The question of whether the genetic associations between PTSD and alcohol consumption differs compared to AUD has not been explicitly studied, to our knowledge. However, work on other psychiatric phenotypes suggests that for some, like smoking behaviors, the genetic correlations between mild and more problematic versions of the phenotype (e.g., cigarettes per day [CPD], nicotine dependence [ND]) are strongly positively correlated with each other (rG=.95; Quach et al., 2020). Additionally, their correlations with other disorders (e.g., schizophrenia) are in the same direction (e.g., both positive) but of varying sizes (Hartz et al., 2018). Research on major depression, which is closely related to PTSD, found positive genetic correlations between major depression and AD and alcohol quantity, but negative genetic correlations between major depression and alcohol frequency (Polimanti et al., 2019). Thus, we aim to test whether using alcohol consumption -related phenotypes yields similar estimates to problem alcohol-related phenotypes.

The current study adds to this literature by estimating genetic correlations from GWASs summary statistics for PTSD (as well as re-experiencing symptoms), and a range of alcohol phenotypes. The latter include drinks per week (DPW), AUDIT-C (alcohol frequency, quantity, and frequency of 6+ drinks), problems (P) score from the AUDIT (AUDIT-P; including 7 items assessing problems; e.g., unable to stop drinking once you started), as well as total score (AUDIT-T; comprised of AUDIT-C and -P), maximum alcohol intake (typical habitual daily maximum usage), AUD (using DSM-V diagnosis), and AD (using DSM-IV diagnosis). In so doing, it adds to previous work by examining not only the genetic association between PTSD and AUD-related outcomes, but also PTSD and other alcohol phenotypes including more normative use, which has been generally neglected in the PTSD-alcohol comorbidity literature with few exceptions (Mallard et al., 2021). Finally, this study attempts to examine whether findings are consistent between those of European Ancestry (EA) and African Ancestry (AA) individuals—the latter of which is particularly important given the lack of diversity in genomic studies (Bentley, Callier, & Rotimi, 2017; Peterson et al., 2019; Sirugo, Williams, & Tishkoff, 2019). This study leverages large-scale GWASs summary statistics from a number of consortia (i.e., Psychiatric Genomics Consortium [PGC]-PTSD and Substance Use Disorder (SUD)-AD, United Kingdom Biobank [UKB], Million Veterans Program [MVP], 23andMe, and GWAS & Sequencing Consortia of Alcohol and Nicotine Use [GSCAN]).

## Methods

### Samples

#### PTSD Samples and Phenotypes

PTSD case/control status came from the PGC-PTSD Freeze 2 dataset (PTSD), which consists of over 50 separate datasets plus the UKB (Nievergelt et al., 2019). In analyses utilizing alcohol use data from the UKB, PGC-PTSD PTSD case status reflects primarily lifetime PTSD diagnosis, but also includes current diagnosis when lifetime was not available (30 of 57 cohorts in Freeze 2 provided lifetime data). PGC-PTSD case/control status data were available for both EA and AA samples. PGC-PTSD Freeze 2 were used instead of Freeze 1.5 because of the increase in sample size and inclusion of AA individuals (EA: Total PTSDf1.5 N = 48,471; PTSDf2.0 N = 174,659; AA: Total PTSDf2.0 N = 15,339) (Nievergelt et al., 2019).

Two PTSD-related variables were used: DSM-based PTSD and a PTSD re-experiencing score. PTSD re-experiencing symptoms (PTSD Re-Exp) came from an assessment of the PTSD Checklist (PCL) DSM-IV version (Wilkins, Lang, & Norman, 2011) in the MVP (Gelernter, Sun, Polimanti, Pietrzak, Levey, Bryois, et al., 2019), selected as it is the symptom cluster most distinctive for PTSD compared to other disorders. This sample and phenotype contributed to both EA (N = 146,660) and AA (N = 19,983) analyses.

#### Alcohol Samples and Phenotypes

AUD and AD GWAS summary statistics were available in two datasets. AD case/control data came from the PGC-SUD (Walters et al., 2018). Cases were defined as meeting criteria for a DSM-IV (DSM-III-R for one study) diagnosis of AD and all controls were alcohol exposed. The PGC-SUD AD phenotype contributed to both EA (N = 46,568) and AA (N = 6,280) analyses. AUD case/control status was used from the MVP dataset, defined as ICD-9 or ICD-10 codes for dependence or abuse as obtained from the Veteran’s Affairs electronic health records (EHR); participants with at least one inpatient or two outpatient alcohol-related ICD-9/10 codes (from 2000-2018) were considered AUD cases (Kranzler et al., 2019). AUD case/control status in MVP is available for EA (N = 267,391) and AA (N = 56,648) samples.

Alcohol consumption-related GWASs summary statistics were available for a number of phenotypes. Specifically, a measure of average DPW came from the GSCAN consortium and the UKB (Liu et al., 2019) available in EA samples only. DPW was defined as the average number of drinks a participant reported drinking each week, aggregated across all types of alcohol. In studies that reported binned response ranges (e.g., 1-4 drinks), the midpoint of the range was used (Liu et al., 2019). Summary statistics for DPW within UKB and GSCAN were examined combined (N = 941,280) as well as separately (GSCAN: N = 526,937; UKB: N = 414,343). The AUDIT (Saunders, Aasland, Babor, De la Fuente, & Grant, 1993) was available in multiple forms and studies. General consumption was measured using the AUDIT-C subscale, which consists of the first three items of the AUDIT and measures past-year typical quantity and frequency of drinking as well as one item measuring frequency of heavy/binge drinking (Bush, Kivlahan, McDonell, Fihn, & Bradley, 1998). AUDIT-C data were available in two datasets, from the EHR data of the annual AUDIT-C assessment in MVP from 2007-2017 (Kranzler et al., 2019) and as part of the full AUDIT assessment in an online follow-up of the UKB (Sanchez-Roige et al., 2019). AUDIT-C data were available in MVP for both EA (206,254) and AA (56,495) ancestries and EA only in UKB (N = 121,604). The full AUDIT score (i.e., AUDIT-T) was also available in the 23andMe and UKB datasets (Sanchez-Roige et al., 2019) in EA samples (23andMe: N = 20,328; UKB: N =121,604). The AUDIT-P scale, the score on items 4-10 of the AUDIT, which focuses on the problematic consequences of drinking, was used from the UKB in EA samples (N = 121,604). Finally, in the MVP data, a quantitative measure of maximum habitual alcohol consumption in a typical month (Max. Alc.; Gelernter, Sun, Polimanti, Pietrzak, Levey, Lu, Hu, Li, Radhakrishnan, & Aslan, 2019) was used as a measure of more problematic consumption, to reflect typical/habitual maximum usage as opposed to maximum on a single occasion (N = 126,936 for EA, and N = 17,029 for AA).

#### Case/Control Designs

Unbalanced ascertainment in case/control designs can introduce bias in studies using meta-analytic data. Effective sample sizes (Neffs) can help to reduce potential bias in this situation. In this study, we used Neff=4/[(1/ncase)□+□(1/ncontrol)] to calculate the Neff for each phenotype, and used the per-SNP Neffs when available in the summary statistics (e.g., PTSD). This approach takes in account the impact of potential bias and reduced power introduced by unbalanced ascertainment of the number of cases and controls across cohorts analyzed under a liability scale (e.g., Walters et al., 2018; Hang Zhou et al., 2020).

#### Genotyping, Quality Control, and Imputation

The existing summary statistics used in the present analyses have previously gone through quality-control pipelines used for the specific consortia (e.g., PGC quality control pipeline including filtering to remove SNPs with imputation information value < .90 and MAF < .01; Sullivan, 2010). The analytic pipeline for the present analyses incorporates further filtering processes including removal of SNPs based on a minimum Neff and per-SNP sample variation (e.g., SNP filtering keeping variants within at least 80% of the total Neff) and variants that are either not SNPs or are strand-ambiguous.

#### SNP heritability and Genetic Correlation Analysis

Analyses of SNP-based heritability (h^2^_SNP_) and genetic correlation (r_G_) were conducted using the cross-trait linkage disequilibrium (LD) score regression approach and LD score regression software (LDSC) (Bulik-Sullivan et al., 2015; open-source LDSC pipeline, version 1.0.1, github.com/bulik/ldsc; =Bulik-Sullivan et al., 2015) which requires GWAS summary statistics in samples of unrelated individuals. The approach estimates r_G_s by replacing the χ^2^ with the *z* scores from both studies and the genetic covariance is then estimated using the slope from the regression of both *z* scores on LD scores. Normalizing genetic covariance by h^2^_SNP_ yields the genetic correlation. Multiple testing was adjusted using false discovery rate (FDR) correction.

Because LDSC requires single ancestry summary statistics as input, analyses were conducted separately for EA and AA samples (see *Supplementary Table 1*). For the EA samples, precomputed LD scores came from the 1000 Genomes Project Europeans (https://data.broadinstitute.org/alkesgroup/LDSCORE/eur_w_ld_chr.tar.bz2). For the AA samples, the AA specific LD scores (subset under UKBB.AFR prefix) from the UKB pan-ancestry LD scores (Pan-UKB team; https://pan.ukbb.broadinstitute.org, 2020) were used.

#### Tissue Enrichment Analysis

At the tissue level, data from 53 human tissues (Genotype-Tissue Expression [GTEx; https://gtexportal.org/home/] project, version 7)(Lonsdale et al., 2013) were used. Post hoc analyses were performed using the partitioned LD score regression software at the default settings (tissues were provided to GTEx by LDSC software).

## Results

### Heritability

Liability scale h^2^_SNP_ statistics were estimated for case/control phenotypes (i.e., PTSDf2, AUD and AD) in EA and AA summary statistics. *Table 1* shows the computed h^2^_SNP_ estimates for those of EA and AA from GWASs used for the analyses below.

**Table 1.** Computed SNP-based Heritability of PTSD and Alcohol Phenotypes – EA and AA

| Phenotype | Data Source (n) | Scale | Sample Prev. | Population Prev. | $h^2_{\text{SNP}}$ | SE | Estimate from Original Publication |
| --- | --- | --- | --- | --- | --- | --- | --- |
| European Ancestry (EA) |  |  |  |  |  |  |  |
| DPW | GSCAN and UKB (n=941,280) | Obs. | - | - | 0.048 | 0.002 | Not Reported ((Liu et al., 2019) |
| DPW | GSCAN (n=526,937) | Obs. | - | - | 0.040 | 0.003 | 0.042[.002] (Liu et al., 2019) |
| DPW | UKB (n=414,343) | Obs. | - | - | 0.066 | 0.004 | 0.74[0.009], 0.79[0.004](Liu et al., 2019) |
| AUDIT-C | MVP (n=206,254) | Obs. | - | - | 0.068 | 0.005 | 0.068[0.005](Kranzler et al., 2019) |
| AUDIT-C | UKB (n=121,604) | Obs. | - | - | 0.084 | 0.006 | 0.084[0.0055](Sanchez-Roige et al., 2019) |
| AUDIT-T | 23andMe (n=20,328) | Obs. | - | - | 0.089 | 0.025 | Not reported (Sanchez-Roige et al., 2019) |
| AUDIT-T | UKB (n=121,604) | Obs. | - | - | 0.059* | 0.005 | 0.086[0.005](Sanchez-Roige et al., 2019) |
| Max. Alc. | MVP (n=126,938) | Obs. | - | - | 0.078 | 0.006 | 0.078[(Gelernter...Lu et al., 2019) |
| AUDIT-P | UKB (n=121,604) | Obs. | - | - | 0.059 | 0.005 | 0.059[0.048]( Sanchez-Roige et al., 2019) |
| AUD | MVP1n2 (n=267,391) | Liab. | 0.183 | 0.159 | 0.108* | 0.006 | 0.056[0.004](Kranzler et al., 2019) |
| AD | PGC (n=46,568) | Liab. | 0.248 | 0.159 | 0.093 | 0.021 | 0.090[0.019](Walters et al., 2018) |
| PTSD Re-Exp | MVP (n=146,660) | Obs. | - | - | 0.068 | 0.005 | 0.067[SE:.005]<br>(Gelernter...Bryois et al., 2019) |
| PTSD | PGC (n=174,659) | Liab. | 0.157 | 0.100 | 0.082* | 0.015 | 0.04[95%CI: 0.02-0.05]<br>(Nievergelt et al., 2019) |
| African Ancestry (AA) |  |  |  |  |  |  |  |
| AUDIT-C | MVP (n=56,495) | Obs. | - | - | 0.061 | 0.016 | 0.062[0.016](Kranzler et al., 2019) |
| Max. Alc. | MVP (n=17,029) | Obs. | - | - | 0.086 | 0.041 | Not Reported (Gelernter...Lu et al., 2019) |
| AUD | MVP (n=56,648) | Liab. | 0.305 | 0.159 | 0.136 | 0.033 | 0.100[0.022](Kranzler et al., 2019) |
| AD | PGC (n=6,280) | Liab. | 0.531 | 0.111 | 0.277 | 0.164 | Unstable/Not Reported (Walters et al., 2018) |
| PTSD Re-Exp | MVP (n=19,983) | Obs. | - | - | 0.067 | 0.041 | 0.048[SE:0.039]<br>(Gelernter...Bryois et al., 2019) |
| PTSD | PGC (n=15,339) | Liab. | 0.284 | 0.100 | 0.193* | 0.072 | 0.02[95%CI:-0.04-0.09]<br>(Nievergelt et al., 2019; note: this was using GCTA not LDSC) |

Note: SNP=Single Nucleotide Polymorphism, PTSD=Posttraumatic Stress Disorder, EA=European ancestry, AA=African ancestry, $h^2$ =Heritability, SE=standard error, Re-Exp=reexperiencing, MVP= Million Veteran Program, AD=Alcohol Dependence, AUD=Alcohol Use Disorder, Max. Alc.=Maximum Alcohol Intake, AUDIT=Alcohol Use Disorder Identification Test, T=total, P=problems, C=consumption, DPW=Drinks per Week, UKB=UK Biobank, GSCAN= GWAS & Sequencing Consortium of Alcohol and Nicotine Use, PGC=Psychiatric Genomics Consortium, Liab.=Liability, Obs.=Observed, Prev.=Prevalence. Sample and population prevalence were used for analyses including binary traits to correct for ascertainment bias. $h^2_{\text{SNP}}$ estimates in this study were computed using LDSC software and the corresponding GWAS summary statistics. For estimates from the original publications, SEs are included when provided in text and when unavailable, 95% CIs were included. When neither was provided in the original manuscript, neither is presented in the table. Four $h^2_{\text{SNP}}$ estimates (noted with an asterisk) are different than those from the original publications. The differences are likely due to additional filtering (see *Method* section), in addition to effective sample sizes calculations (see *Method* section) that take in account differences in ascertainment of unbalanced cohorts in the case of the three $h^2_{\text{SNP}}$ estimates under the liability scale with an asterisk.

### Genetic Correlations

We estimated the r_G_ s of PTSD phenotypes (i.e., PTSD [PGC], and PTSD Re-Exp [MVP]) with DPW (from the GSCAN, UKB and both combined), AUDIT-C (from the MVP and UKB), AUDIT-T (from 23andMe and UKB), Max. Alc. (from the MVP), AUDIT-P (from the UKB), AUD (from the MVP) and AD (from the PGC) in individuals from EA (see *Table 2* and *Figure 1*). Similarly, we estimated the r_G_s of PTSD and PTSD Re-Exp with AUDIT-C (from the MVP), Max. Alc. (from the MVP), AUD (from the MVP), AD (from the PGC) phenotypes in AA samples (see *Table 3* and *Figure 2*).

**Table 2.**
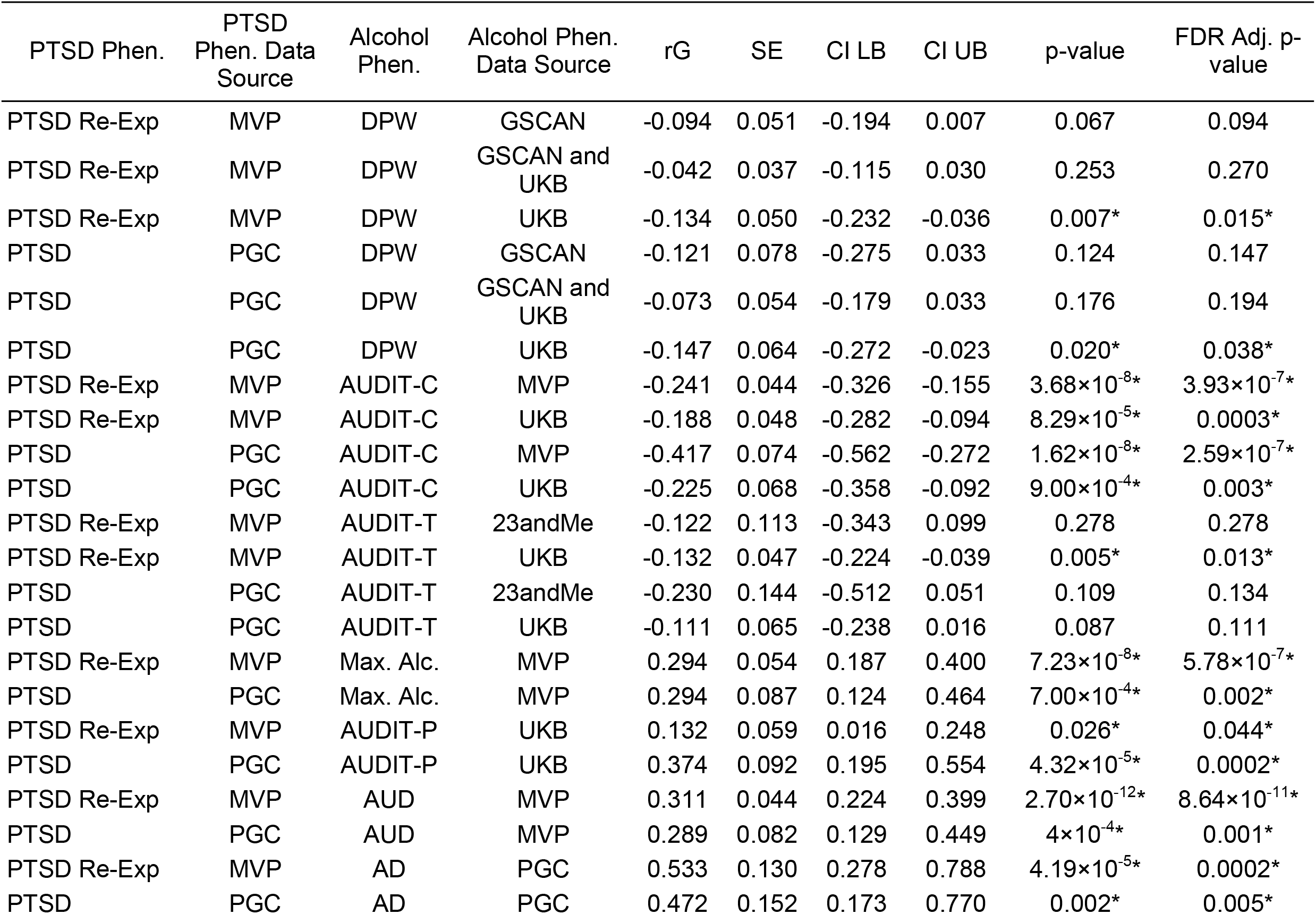
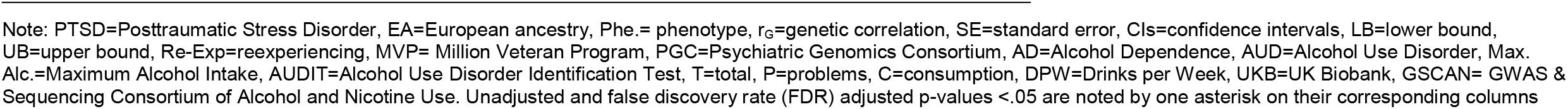
Genetic Correlations of PTSD and Alcohol Phenotypes in people of EA ancestry

**Figure 1.**
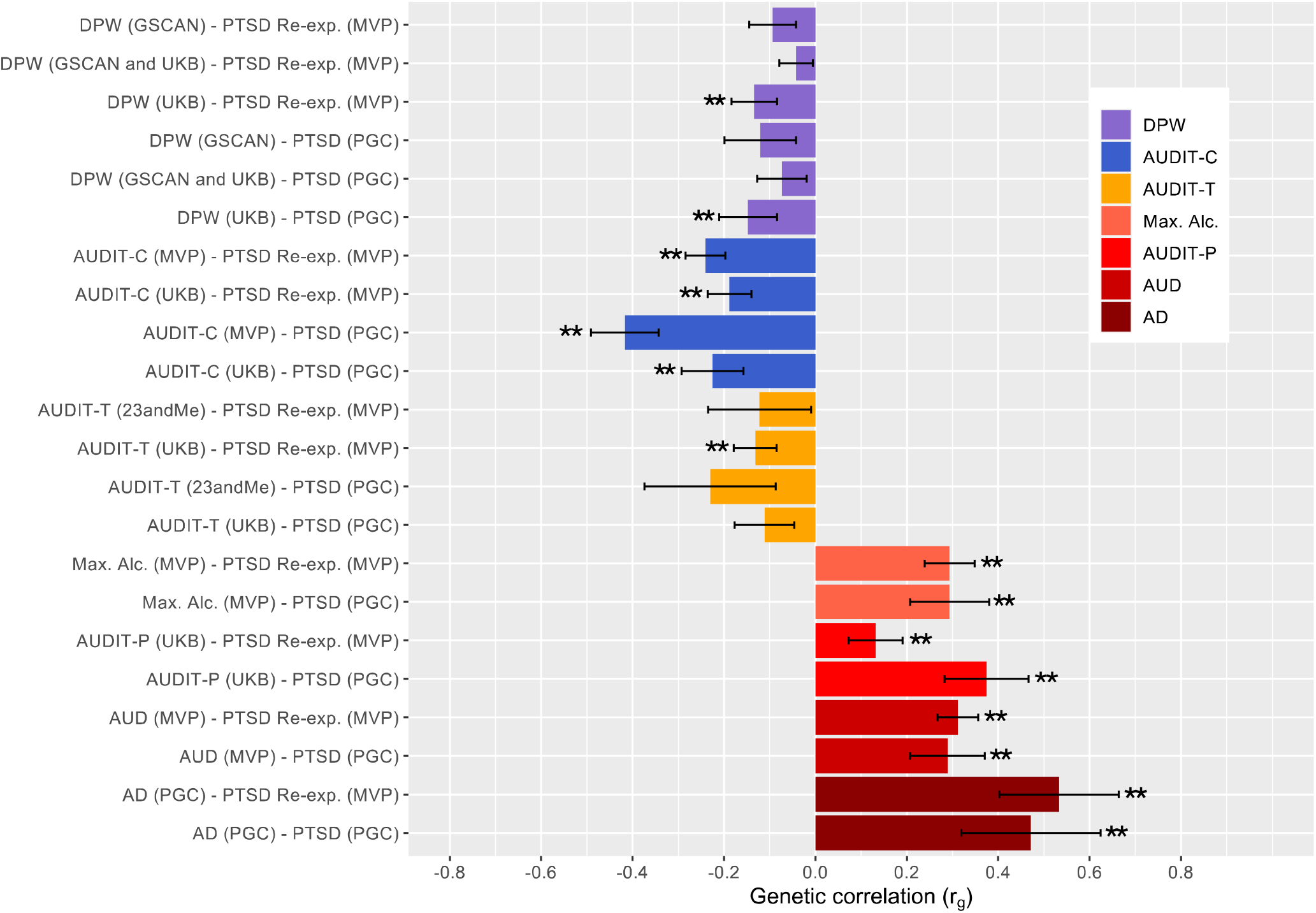
Genetic Correlations of PTSD and Alcohol Phenotypes (+/- SE bars) – EA Note: Unadjusted significant p-values (p<.05) for r_G_s are noted with an asterisk. Those passing FDR adjustment are noted with an additional asterisks (total of two asterisks for those passing FDR adjustment). PTSD=Posttraumatic Stress Disorder, EA=European ancestry, SE=standard error, DPW=Drinks per Week, GSCAN= GWAS & Sequencing Consortium of Alcohol and Nicotine Use, UKB=UK Biobank, AUDIT=Alcohol Use Disorder Identification Test, T=total, P=problems, C=consumption, Max. Alc.=Maximum Alcohol Intake, AUD=Alcohol Use Disorder, AD=Alcohol Dependence, PGC=Psychiatric Genomics Consortium, Re-exp.=reexperiencing, MVP= Million Veteran Program. Alcohol phenotypes are ordered from more typical to more problematic (top to bottom) and color coded by each type of phenotype (i.e., DPW, AUDIT-C, AUDIT-T, Max. Alc., AUDIT-P, AUD, AD) to draw attention to difference in findings.

**Table 3.** Genetic Correlations of PTSD and Alcohol Phenotypes in people of AA ancestry

| PTSD Phe. | PTSD Phe. Data Source | Alcohol Phe. | Alcohol Phe. Data Source | rG | SE | CI LB | CI UB | p-value | FDR Adj. p-value |
| --- | --- | --- | --- | --- | --- | --- | --- | --- | --- |
| PTSD | PGC | AUDIT-C | MVP | 0.145 | 0.209 | -0.265 | 0.554 | 0.489 | 0.719 |
| PTSD Re-Exp | MVP | AUDIT-C | MVP | 0.699 | 0.328 | 0.057 | 1.340 | 0.033* | 0.264 |
| PTSD | PGC | Max. Alc. | MVP | -0.123 | 0.342 | -0.794 | 0.548 | 0.719 | 0.719 |
| PTSD Re-Exp | MVP | Max. Alc. | MVP | -0.275 | 0.504 | -1.260 | 0.713 | 0.586 | 0.719 |
| PTSD | PGC | AUD | MVP | 0.182 | 0.185 | -0.180 | 0.544 | 0.325 | 0.719 |
| PTSD Re-Exp | MVP | AUD | MVP | 0.266 | 0.270 | -0.263 | 0.796 | 0.324 | 0.719 |
| PTSD | PGC | AD | PGC | 0.233 | 0.305 | -0.365 | 0.832 | 0.445 | 0.719 |
| PTSD Re-Exp | MVP | AD | PGC | 0.195 | 0.522 | -0.829 | 1.220 | 0.709 | 0.719 |

Note: PTSD=Posttraumatic Stress Disorder, Phe.= phenotype, rG=genetic correlation, SE=standard error, CIs=confidence intervals, LB=lower bound, UB=upper bound, Re-Exp=reexperiencing, MVP=Million Veteran Program, PGC=Psychiatric Genomics Consortium, AD=Alcohol Dependence, AUD=Alcohol Use Disorder, Max. Alc.=Maximum Alcohol Intake, AUDIT=Alcohol Use Disorder Identification Test, C=consumption. The p-value <.05 is noted by one asterisk. None passed FDR adjustment.

**Figure 2.**
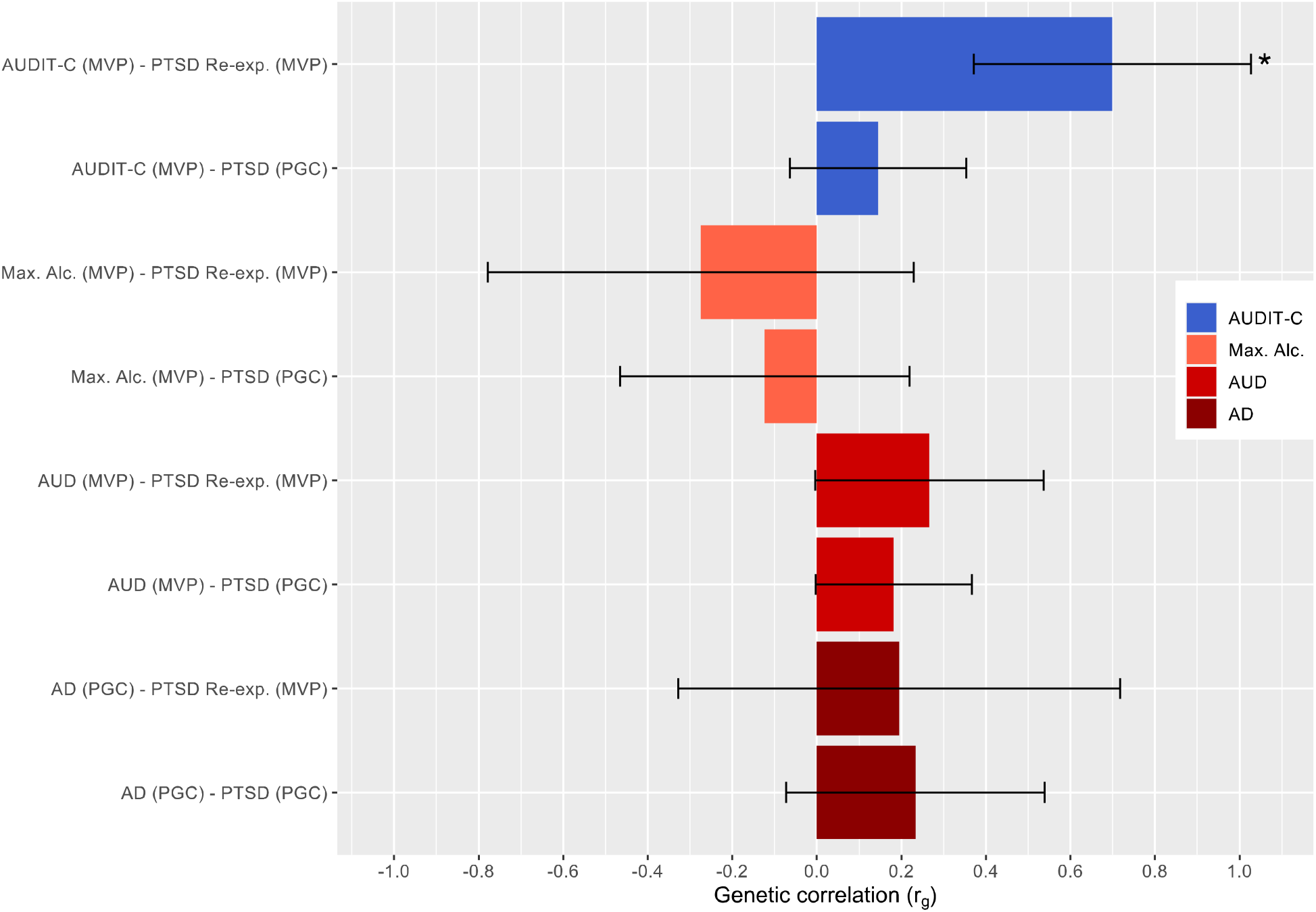
Genetic Correlations of PTSD and Alcohol Phenotypes (+/- SE bars) – AA Note: Unadjusted significant p-values (p<.05) for r_G_s are noted with an asterisk. Those passing FDR adjustment are noted with an additional asterisks (total of two asterisks for those passing FDR adjustment). PTSD=Posttraumatic Stress Disorder, AA=African ancestry, SE=standard error, AUDIT=Alcohol Use Disorder Identification Test, C=consumption, Max. Alc.=Maximum Alcohol Intake, AUD=Alcohol Use Disorder, AD=Alcohol Dependence, PGC=Psychiatric Genomics Consortium, Re-exp.=reexperiencing, MVP= Million Veteran Program. Alcohol phenotypes are ordered from more typical to more problematic (top to bottom) and color coded by each type of phenotype (i.e., AUDIT-C, Max. Alc., AUD, AD) to draw attention to difference in findings.

#### PTSD-AUD-Related Phenotypes

Notably, the r_G_s between all PTSD phenotypes and AUD-related phenotypes are positive for EA individuals (r_G_s: .132-.533, all FDR adjusted p<.05). The r_G_ estimates for PTSD with Max. Alc., AUDIT-P, AUD and AD phenotypes were positive and moderate (r_G_s: .289-.533, all FDR adj. p<.01) with the exception of AUDIT-P and PTSD Re-Exp, which was small, still positive (r_G_: .132, FDR adj. p=.044). All these r_G_ estimates passed FDR adjustment. Notably, the highest r_G_s were between the two PTSD phenotypes and AD (r_G_s: .472-.533, p<.001).

A similar trend of positive r_G_s was observed for PTSD phenotypes (i.e., PTSD, PTSD Re-Exp) with AUD and AD phenotypes on AA samples (r_G_s: .182-.266, NS). Conversely, PTSD phenotypes and Max. Alc. correlated negatively for individuals of this ancestry (r_G_: -.123-.275, NS). However, regardless of r_G_ direction, these estimates yielded relatively large standard errors (SEs) and non-significant results (*p*>.05).

#### PTSD-Alcohol Consumption-Related Phenotypes

The r_G_s between the PTSD phenotypes and alcohol consumption phenotypes (i.e., DPW, AUDIT-C, AUDIT-T) are negative and varying in degree from small to large for those of AA (AUDIT-C and PTSD Re-Exp r_G_: .145, NS; AUDIT-C and PTSD r_G:_ .699, unadjusted p<.05), in contrast to the positive and mostly moderate r_G_s of PTSD and AUD phenotypes. For those of EA, the genetic correlations between PTSD and PTSD Re-Exp and DPW across samples were negative, small, and non-significant (*p*>.05), with the exception of DPW (UKB) genetic correlations with both PTSD phenotypes, passing FDR adjustment. The r_G_s between PTSD phenotypes with AUDIT-C and -T were low to moderate. However, the r_G_s with AUDIT-C across samples were moderate (r_G_s: -.225 to -.417; with the exception of AUDIT-C [UKB] - PTSD Re-exp r_G_s: -.188), significant and passed FDR correction. Whereas the genetic correlation between AUDIT-T (UKB) and PTSD Re-exp was the only significant association, albeit small, among all the AUDIT-T analyses for EA. The highest r_G_ among PTSD and alcohol consumption-related phenotypes was that of AUDIT-C (MVP) with PTSD (r_G_: -.417, FDR adj. p=2.59×10^−7^).

The r_G_s using samples of AA individuals for PTSD phenotypes (i.e., PTSDf2, PTSD Re-Exp) and AUDIT-C were positive, and only that with PTSD Re-Exp was significant; although it did not pass FDR adjustment. Notably, this positive r_G_ estimate in AA individuals, contrasts with the negative r_G_s estimates in EA samples for the same phenotypes. See *Supplementary Figure 1* for boxplot with whiskers display of genetic correlations for those of EA and AA.

### Post-Hoc Tissue Enrichment

In EA samples, only the GWAS from MVP PTSD Re-experiencing symptoms, Drinks Per Week (GSCAN and UKB) and Drinks Per Week (UKB) met the FDR significance threshold for specific tissue enrichment (see *Supplementary Figure 2)*. These three GWAS that met for this threshold exceeded the FDR significance value for only tissues having to do with the brain (i.e., not other tissues). In AA samples, none of the included GWAS met the FDR threshold for significance for tissue enrichment (see *Supplementary Figure 3*).

## Discussion

PTSD commonly co-occurs with increased alcohol consumption and AUD. While our previous work has demonstrated that there is a molecular genetic correlation between PTSD and AUD (Sheerin et al., 2020), the goal of this study was to determine whether the genetic correlations with PTSD extend to other dimensions of more typical alcohol consumption, and if the architecture of the genetic association with PTSD differs for alcohol consumption and AUD. Further, we aimed to test these associations using both EA and AA summary statistics using data from the latest GWASs of PTSD, alcohol consumption-related, and AUD-related phenotypes. Among EA analyses, this study found positive and significant genetic correlations between PTSD and AUD-related phenotypes, whereas negative with a few non-significant genetic correlations observed for PTSD and alcohol consumption-related phenotypes. These results indicate that which alcohol phenotype one uses in analyses absolutely matters, and that alcohol use is certainly not “close enough” as a proxy for AUD in examining its genetic associations with other conditions. Among those of AA, potentially due to having reduced power, associations were generally non-significant, with the exception of a positive correlation (not passing FDR adjustment) between PTSD Re-Exp symptoms and AUDIT-C, which was not observed for EA individuals.

PTSD, alcohol consumption, and AUD have been shown to be heritable in both twin (Heath, Jardine, & Martin, 1989; Kaprio et al., 1987; Knopik et al., 2004; Stein, Jang, Taylor, Vernon, & Livesley, 2002) and molecular-genetic studies (Clarke et al., 2017; Sanchez-Roige et al., 2019; Stein et al., 2016). Not surprisingly, the h^2^_SNP_ estimates from our study are smaller than those from twin studies, finding estimates of ∼.38 for PTSD, .36-.40 for consumption, and .47 for alcohol misuse, with overall similarities among EA and AA participants. Although the estimated heritability for PTSD ((h^2^_SNP_ =19.3% vs. 8.2%) and Alcohol Dependence (27.7% vs. 9.3%) appear higher in AA compared to EA respectively they are not statistically different as there are large standard errors on the AA estimates (see Table 1). This is likely an artifact of the relatively greater number of participants of EA included in the discovery GWAS datasets. Large genome-wide studies have historically focused on participants of EA, leading to an important gap in knowledge regarding genetic epidemiology across diverse ancestral groups that our field must address.

Consistent with previous work (Sartor et al., 2011; Sheerin et al., 2020), this study found positive genetic correlations between PTSD and AUD-related phenotypes (Max. Alc., AUDIT-P, AUD, AD) among individuals of EA. Our findings are also consistent with a recent paper finding a moderate positive genetic correlation between PTSD and problematic alcohol use (rG=.49), and a more modest genetic correlation between PTSD and the specific portion of problematic alcohol use unique from a larger externalizing factor (rG=.26) (Barr et al., 2021). However, when investigating genetic correlations between PTSD and alcohol consumption-related phenotypes, findings generally suggested negative and significant (e.g., in the case of DPW correlations using UKB data) or negative and non-significant (e.g., in the case of DPW correlations using GSCAN) associations with PTSD. These discrepancies may have arisen because of differing sample characteristics or differing numbers of studies contributing to these statistics. However, in general, the genetic associations between PTSD and alcohol consumption phenotypes were different from those of PTSD-AUD.

These results suggest that different genetic factors may exist for individuals with PTSD and increased alcohol consumption and for individuals with comorbid PTSD and AUD. These results are also consistent with the very small amount of work conducted examining genetic associations between PTSD and alcohol use. Specifically, a paper by our group employing Mendelian Randomization (MR) as the primary method also found in secondary analyses using LDSC a non-significant genetic association between PTSD and DPW among those of EA (Bountress et al., 2021). Additionally, a similar trend has been observed with other psychiatric disorders in terms of the genetic association between alcohol consumption vs. disorder, specifically major depressive disorder, which has substantial genetic overlap with PTSD (Polimanti et al., 2019; Sanchez-Roige et al., 2019; Walters et al., 2018; Zhou et al., 2020). These authors observed positive genetic correlations between major depression and alcohol dependence. However, they also observed negative genetic correlations between major depression and frequency of alcohol consumption. Further, the same trend in attention-deficit/hyperactivity disorder (ADHD) was observed such that problematic drinking was positively genetically correlated with ADHD and alcohol consumption was negatively genetically correlated with ADHD (Sanchez-Roige et al., 2019). Future work might benefit from additional MR analyses examining the potential causal relations between PTSD and more alcohol phenotypes.

Interestingly, one study found that the genetic association between AUDIT-C and AD was initially negative, but became positive when the “healthy volunteer” effect that tends to occur in alcohol frequency data was taken into account (Mallard et al., 2021). Thus, in cases where the associations between alcohol consumption frequency and alcohol consumption quantity and other phenotypes are in opposing directions, this may be because the frequency item is positively genetically correlated with high SES (Mallard & Sanchez-Roige, 2021; Marees et al., 2020). Another potential explanation for the negative association between consumption and other psychopathology-related outcomes may be related to the finding that some with greater disease burden have in turn, reduced or limited their alcohol consumption (Xue et al., 2020). Our study is the first to observe a negative genetic correlation between PTSD and alcohol consumption-related phenotypes, among those of EA. Findings from this study, together with the previous literature on genetic correlations between other psychiatric disorders and problematic vs. typical alcohol use, consistently indicate that problematic alcohol use and more typical alcohol use are genetically associated with other psychiatric disorders in opposite directions. These findings suggest that alcohol-related behaviors are heterogenous, and specifically that the genetic associations between consumption and problem alcohol phenotypes and PTSD differ in strength and direction.

Importantly, we note that this pattern of results was driven by participants of EA, and that one contrasting finding was observed among AA participants. This difference in finding may be due to discrepancy in size of the discovery GWAS, or it may reflect social-cultural differences regarding alcohol use patterns among African Americans and Whites in the US. For example, among individuals who self-identify as Black or African American, regular and heavy alcohol use is much less common compared to Whites (Delker, Brown, & Hasin, 2016). Increasing diversity in large genetic studies will enable improved understanding of the epidemiology of PTSD, alcohol use behaviors, and their comorbidity. We also note that although prior work by our group found sex differences in the genetic associations between PTSD and AD (Sheerin et al., 2020), we were unable to test whether that difference extended to other alcohol phenotypes, as summary statistics stratified by sex were not available. Future research ought to attempt to test this question.

In terms of the post-hoc enrichment analyses, the finding that for EAs, the GWAS that reached FDR significance all met for brain tissues, but not others, is generally consistent with prior work for PTSD and other neuropsychiatric traits (Dalvie et al., 2021; Gelernter et al., 2019). However, additional work is needed to better understand why only three of the GWAS (MVP PTSD Re-Experiencing, DPW GSCAN, DPW UKB) exceeded the FDR cut-off. For AAs, no GWAS exceeded the FDR threshold. It is possible that this lack of significant effects is due to the smaller n’s among the AA GWAS.

In conclusion, findings from this study extend knowledge regarding the genetic associations of PTSD and AUD, to include a spectrum of alcohol use phenotypes including more typical alcohol use in an ancestrally diverse population. These findings indicated positive genetic associations between PTSD and AUD-related phenotypes and negative genetic correlations between PTSD and alcohol consumption-related phenotypes. Thus, the genetic factors leading to the development of PTSD when considering alcohol consumption are quite different from the those leading to the development of comorbid PTSD-AUD. These findings support the growing number of studies demonstrating the important differences regarding risk factors for alcohol use vs. disorder, and their associations with other psychiatric disorders (e.g., depression, ADHD).

## Data Availability

All data produced/generated in the present study are available upon reasonable request to the authors. However, individual summary statistic files that are referred to may only be obtained by contacting the accompanying consortia/group (e.g., MVP).

---

**Supplementary Figure 1.**
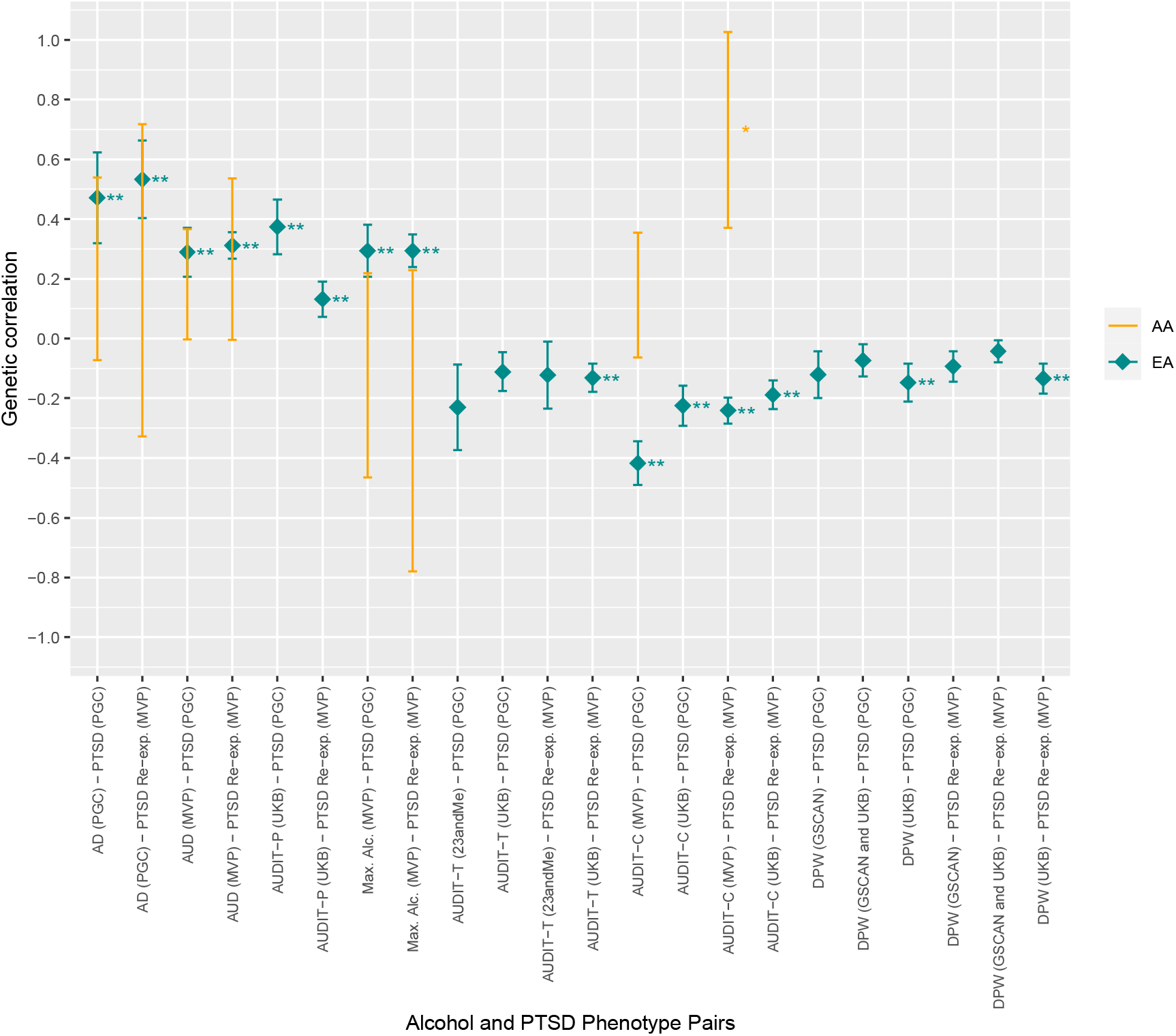
Comparisons of PTSD and Alcohol Phenotypes (+/- SE bars) Genetic Correlations – EA and AA Note: Unadjusted significant p-values (p<.05) for r_G_s are noted with an asterisk. Those passing FDR adjustment are noted with an additional asterisks (total of two asterisks for those passing FDR adjustment). PTSD=Posttraumatic Stress Disorder, SE=standard error, EA=European ancestry, AA=African ancestry, DPW=Drinks per Week, GSCAN= GWAS & Sequencing Consortium of Alcohol and Nicotine Use, UKB=UK Biobank, AUDIT=Alcohol Use Disorder Identification Test, T=total, P=problems, C=consumption, Max. Alc.=Maximum Alcohol Intake, AUD=Alcohol Use Disorder, AD=Alcohol Dependence, PGC=Psychiatric Genomics Consortium, Re-exp.=reexperiencing, MVP= Million Veteran Program. Alcohol phenotypes are ordered from more problematic to more typical (left to right). Genetic correlation estimates by ancestry are coded by color and shape.

**Supplemental Figure 2.**
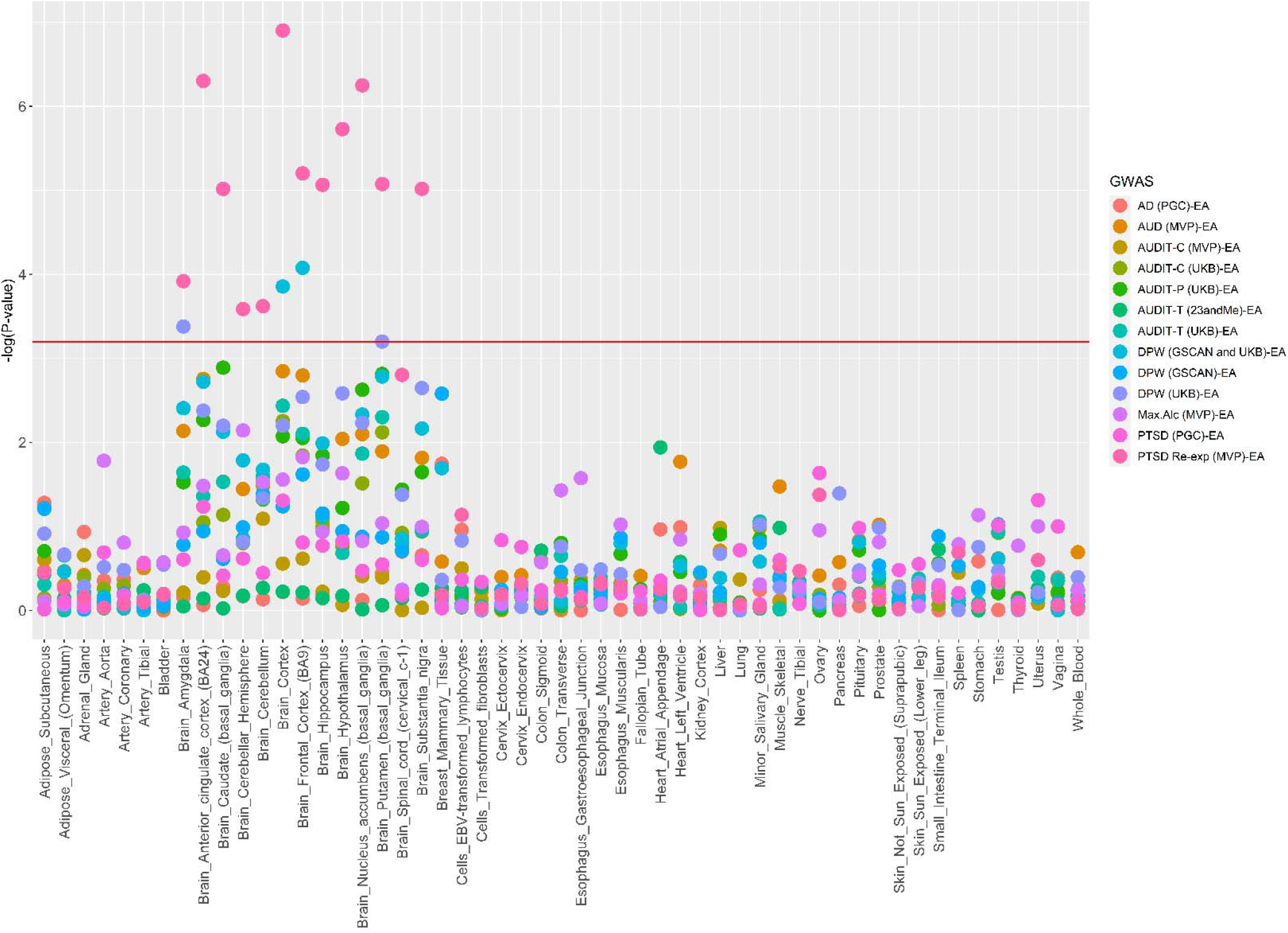
Tissue Enrichment Analyses among European Ancestry (EA) individuals. Note: Red line indicates the FDR significance value, so all points above the line indicate GWAS that met this threshold. Notably, only the GWAS from MVP PTSD Re-experiencing symptoms, Drinks Per Week (GSCAN and UKB) and Drinks Per Week (UKB) met the FDR significance threshold.

**Supplementary Figure 3.**
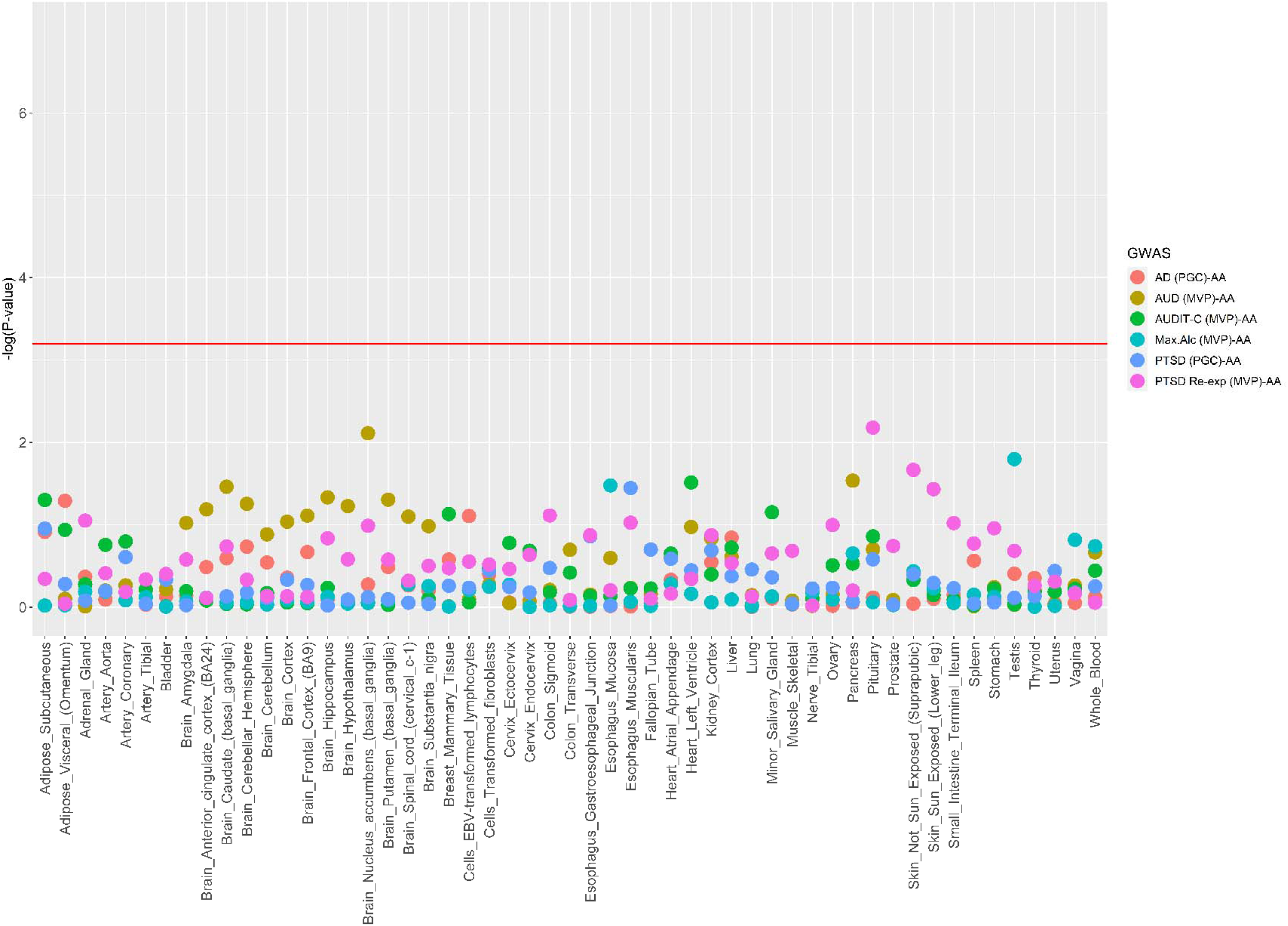
Tissue Enrichment Analyses among African Ancestry (AA) individuals. Note: Red line indicates the FDR significance value. Notably, none of the included GWAS met the FDR threshold for significance.

**Supplementary Table 1.** List of PTSD and alcohol phenotypes summary statistics by ancestry used in this study

| Phenotype | Data Source | Ancestry |
| --- | --- | --- |
| DPW | GSCAN and UKB | EA |
| DPW | GSCAN | EA |
| DPW | UKB | EA |
| AUDIT-C | MVP | EA and AA |
| AUDIT-C | UKB | EA |
| AUDIT-T | 23andMe | EA |
| AUDIT-T | UKB | EA |
| Max. Alc. | MVP | EA and AA |
| AUDIT- P | UKB | EA |
| AUD | MVP | EA and AA |
| AD | PGC | EA and AA |
| PTSD Re-Exp | MVP | EA and AA |
| PTSD | PGC | EA and AA |

Note: PTSD=Posttraumatic Stress Disorder, Re-Exp=reexperiencing, MVP= Million Veteran Program, AD=Alcohol Dependence, PGC= Psychiatric Genomics Consortium, AUD=Alcohol Use Disorder, Max. Alc.=Maximum Alcohol Intake, AUDIT=Alcohol Use Disorder Identification Test, T=total, P=problems, C=consumption, DPW=Drinks per Week, UKB=UK Biobank, GSCAN= GWAS & Sequencing Consortium of Alcohol and Nicotine Use, EA=European ancestry, AA=African ancestry

